# Effect of Cervical Epidural Stimulation After Neurological Damage: Protocol for Phase I/II Open Label Controlled CE-STAND Trial

**DOI:** 10.64898/2025.12.02.25341062

**Authors:** NL Mansour, M Keller-Ross, WC Low, DY Balser, AM Parr

## Abstract

**Introduction:** Spinal cord injury (SCI) affects over 18,000 individuals annually, leading to significant motor and autonomic dysfunctions that reduce quality of life (QoL). Epidural spinal cord stimulation (eSCS) has shown potential in restoring function in chronic individuals with SCI. However, research on cervical, and autonomic dysfunction-related SCI remains limited. This study will assess the safety and efficacy of eSCS in improving autonomic and volitional functions, as well as truncal stability, bowel, bladder, and sexual function, and overall QoL.

**Methods:** The study is a Phase I/II trial in which participants will serve as their own controls. The Abbott Eterna epidural stimulator system will be implanted, and assessments will occur every three months. The study will be conducted over a period of four years. A total of 36 participants will be enrolled and followed for one year.

**Analysis:** Bayesian Optimization will be used to determine the most effective stimulation settings, with 15 settings programmed at the end of each visit.

**Ethics and dissemination:** FDA and UMN IRB approvals have been secured for the study. Interim reports will be made available through peer-reviewed journals, conference presentations, and clinical network reporting.

**Conclusion:** CE-STAND seeks to advance SCI treatment by expanding eSCS applications in chronic cervical SCI, improving patient outcomes, and reducing healthcare costs. Future studies could consider a larger sample size, long term safety, matched-control, and randomization.

**Registration:** The study is registered on ClinicalTrials.gov (NCT06410001).

**Strengths and Limitations:** *Strengths:* This trial investigates eSCS for improvement of autonomic and volitional function in individuals with SCI, an important missing link in the treatment of SCI. A key innovation is the incorporation of microneurography, the gold standard test used to quantify muscle sympathetic nerve activity (MSNA), which is underinvestigated in SCI neuromodulation research. Furthermore, the trial enrolls cervical individuals with SCI, a population that has previously been underrepresented in this research area. The research design is sound, with well-defined outcomes aimed at maximizing methodological quality and regulatory compliance while minimizing bias. The provision of an interim analysis allows for early review of findings, enabling protocol modifications and resolution of study issues, while also guiding future research.

*Limitations:* A main shortcoming of this trial is the extremely small number of subjects, which can lower statistical power and limit generalizability of the results. The study improves the representativeness of individuals with cervical SCI. However, it does not consider variability in injury levels, which can limit the generalizability of findings. The trial follows up participants over one year, measuring short-term effects, but not tracking the long-term safety and efficacy of the intervention. A second challenge is the variability of subjects in response to stimulation, which can lead to variability in optimization of parameters and treatment outcomes. Since there is no treatment for chronic SCI, participants will serve as their own controls which could challenge blinding of participants.

## Introduction

### Background and Rational

Spinal cord injury (SCI) is a significant and persistent public health challenge, with over 18,000 new cases occurring annually ^1,2^. The proportion of cervical injuries has been rising over the past few decades as has the average age at which the SCI occurs. These variables are likely related, with the aging population being more prone to falls, resulting in cervical injury. Indeed, the proportion of SCI related to falls, compared to motor vehicle accidents has also increased ^3^. There is no indication that this increasing rate of cervical injuries has plateaued.

Chronic SCI remains without readily available treatment options beyond rehabilitation and preventing and treating complications. While many groups have focused on therapies for subacute SCI, treatment for chronic SCI remains an unmet need, with an estimated 296,000 people currently living with chronic SCI in the US ^4^. With improvements in modern healthcare and preventative and treatment strategies for complications of SCI, individuals with SCI are expected to live longer with the total years lived with chronic SCI expected to increase. Thus, improving interventions for chronic SCI will not only address an unmet need, but will improve the QoL, morbidity and mortality for this population.

Although motor paralysis is the most obvious effect of SCI, autonomic dysfunction has been shown to affect QoL, morbidity and mortality ^5^. Cost is estimated at $5.8 million over a lifespan ^6^. The severity of autonomic dysfunction varies based on the level of injury, with dysfunction being more common in higher-level injuries. This includes loss of body temperature regulation, bladder and bowel control, sexual function and sensation, as well as the development of autonomic dysreflexia ^7–9^. Dysfunction in blood pressure (BP) regulation has also been shown to directly contribute to decreased cognition in this population ^10,11^.

eSCS has now emerged as a restorative neuromodulation paradigm for SCI autonomic and volitional comorbidities, independently reported across at least three sites including our group here at the University of Minnesota (UMN) ^8,12–14^. However, research on chronic cervical SCI and autonomic dysfunction is underrepresented, necessitating targeted investigations into novel therapies. CE-STAND is an extension of E-STAND, a study investigating the effect of eSCS in individuals with chronic motor complete thoracic SCI. We are expanding our cohort to include the cervical population (C4-C7). CE-STAND participants will complete a very similar protocol to the E-STAND protocol ^15^ with the major differences being cervical injuries (C4-C7) and the focus on autonomic dysfunction, with the stimulator placement at T10-T12 to be able to compare these participants with those from E-STAND. Further, none of these studies utilize the gold-standard technique microneurography to assess sympathetic activity.

Several earlier studies have shown a significant reduction in resting sympathetic tone following SCI, particularly in individuals with high-level lesions ^16,17^. This reduction has critical implications for autonomic dysfunction in this population. Our protocol addresses this limitation by incorporating direct measurement of MSNA and cardiovascular autonomic regulation. This approach aligns with more recent studies demonstrating the applicability of this test in SCI participants ^18–20^ and provides further confidence that we are using a valid and rigorous method for detecting changes in sympathetic function. Further, our work will take this research to the next step to identify if cervical eSCS can increase sympathetic activity in adults with SCI.

### Trial Design

CE-STAND is a phase I/II study representing a single-arm, sham-controlled crossover study. In which participants will serve as their own controls: stimulation ‘on’ can be compared with stimulation ‘off’ within the same follow-up visit. Thus, concerns about cumulative stimulation-induced plasticity over extended periods are not relevant. This approach allows valid within-subject comparisons while minimizing risk to participants.

### Objectives

This manuscript is highly innovative as it focuses on cardiovascular and cognitive dysfunction in chronic, cervical individuals with SCI. The long-term goal is to determine which individuals with SCI will benefit from eSCS therapy, providing support for this technology to be more widely available. Thus, the goal of this project is to determine whether eSCS can provide the same benefits in patients with cervical injuries as those with thoracic injuries. Accordingly, we will:

Aim 1. Evaluate the safety of eSCS in chronic individuals with SCI. We hypothesize that eSCS is a safe intervention when applied to chronic cervical individuals with SCI.

Aim 2. Evaluate the efficacy of eSCS on autonomic function in chronic cervical individuals with SCI, including cardiovascular and cognitive effects, and compare these results to our prior participants with thoracic injuries. We hypothesize that eSCS will be beneficial in restoring BP, cardiovascular function, and cognitive ability dysregulation through tilt table testing. Further, we hypothesize that microneurography will demonstrate an increase in sympathetic function. However, comparison will not be possible since there is no MSNA data for individuals with thoracic SCI.

Aim 3. Evaluate the efficacy of eSCS on other modalities in chronic cervical individuals with SCI including volitional movement and bowel and bladder and compare these results to our prior participants with thoracic injuries. We hypothesize that eSCS will be beneficial in restoring volitional movement in the lower extremities, but not the upper extremities. Further, we hypothesize that we will also see improvements in other modalities such as truncal stability, and bowel and bladder function.

## Methods: Participants, Intervention, and Outcomes

The CE-STAND study is currently in the final stages of the start-up phase. Recruitment has not begun yet, as we are awaiting receipt of the epidural stimulators.

### Study Setting

CE-STAND will take place at the University of Minnesota Medical Center (UMMC), a Level Two Trauma Center. The surgeries will be performed at the University of Minnesota Clinics and Surgery Center (UMN CSC). The study will be conducted in collaboration with the Cardiovascular Rehabilitation Laboratory at the UMN, which will oversee the autonomic function testing.

### Eligibility Criteria

Individuals with chronic, traumatic SCI will be recruited if they meet the following criteria: Inclusion Criteria:

- Twenty two years of age or older
- Able to undergo the informed consent/assent process
- Stable, motor-complete SCI
- Discrete SCI between C4 and C7 (upper extremity weakness)
- ASIA A or B SCI Classification
- Medically stable in the judgment of the PI
- Intact segmental reflexes below the lesion of injury
- Greater than 1 year since initial injury and at least 6 months from any required spinal instrumentation
- Normal or corrected-to-normal vision to accurately perceive visual stimuli on the N-back task screen, and sufficient hearing ability to accurately perceive auditory stimuli during the task
- Willing to attend all scheduled appointments Exclusion Criteria:
- Upper cervical injury
- Diseases and conditions that would increase the morbidity and mortality of this surgery such as cardiopulmonary.
- Individuals who require respiratory support, including ongoing or intermittent use of a ventilator and/or diaphragmatic pacer
- Inability to withhold antiplatelet/anticoagulation agents perioperatively
- Individuals who have history of recurrent autonomic dysreflexia in the judgment of the PI
- Significant dysautonomia that would prohibit rehabilitation or any history of cerebrovascular accident or myocardial infarction associated with autonomic dysreflexia. A single tilt table test with syncope, presyncope, or SBP < 50 or > 200
- Failure to exhibit cardiovascular autonomic dysfunction on tilt table testing, including any signs or symptoms of syncope or presyncope during or after the test, or a systolic blood pressure (SBP) increase or decrease of ≥20 mmHg from baseline
- Other conditions that would make the subject unable to participate in testing/rehabilitation in the judgment of the PI
- Individuals who have participated in another study within the last 12 months in which they have received fluoroscopic or other related radiation exposure
- Current and anticipated need for opioid pain medications or pain that would prevent full participation in the program in the judgment of the PI
- Clinically significant mental illness in the judgment of the PI
- Treatment with botulinum toxin, intrathecal baclofen pump, and antispasmodics will not be permitted unless discussed with and approved by the study PI
- Individuals with a history of significant depression or drug abuse
- Volitional movements present during EMG testing in bilateral lower extremities
- Unhealed spinal fracture
- Presence of significant contracture with loss of greater than two-thirds range of motion
- Presence of pressure ulcers
- Current Pregnancy

### Sample Size

It is very difficult to utilize a power calculation given our prior experience that all participants recovered some volitional movement and those that experienced autonomic dysfunction also benefited. Using a dichotomous model for power calculation, assigning participants as “responders” or “non-responders” based on whether their BP returns to baseline after application of eSCS, we calculated the following: Assuming that 0% of participants will respond to “eSCS off,” and even if only 25% of participants respond to “eSCS on,” with power set at 90%, only 35 participants would be needed to determine a treatment effect. In reality, we expect many more participants to respond based on our prior experience, and we do not expect to have significant drop-out; thus, 36 participants should be an ample number for our eventual goal.

### Recruitment

Participants will be recruited via flyers made available through the Department of Neurosurgery and Physical Medicine & Rehabilitation clinics. We anticipate that these flyers will be widely disseminated throughout the University. We will use our existing contacts within the SCI community and national conferences for word-of-mouth advertising. Further, we will use the registry of our existing trial, E-STAND, to identify and recruit potentially eligible participants.

A trained researcher will contact participants via email and request additional information, such as medical records, including images of the spinal cord taken after their last spinal surgery. Final eligibility will be determined after the screening appointment. Participants who are identified as potentially eligible will be contacted via email or telephone to confirm their interest in participating.

### Participant Timeline

The study will be conducted over four years (see Figure 1). We anticipate that up to 50 individuals will be enrolled to undergo the screening process. Ultimately, 36 individuals will be implanted with the investigational device. Three participants will be enrolled every three months, with each participant followed for one year.

**Figure 1.**
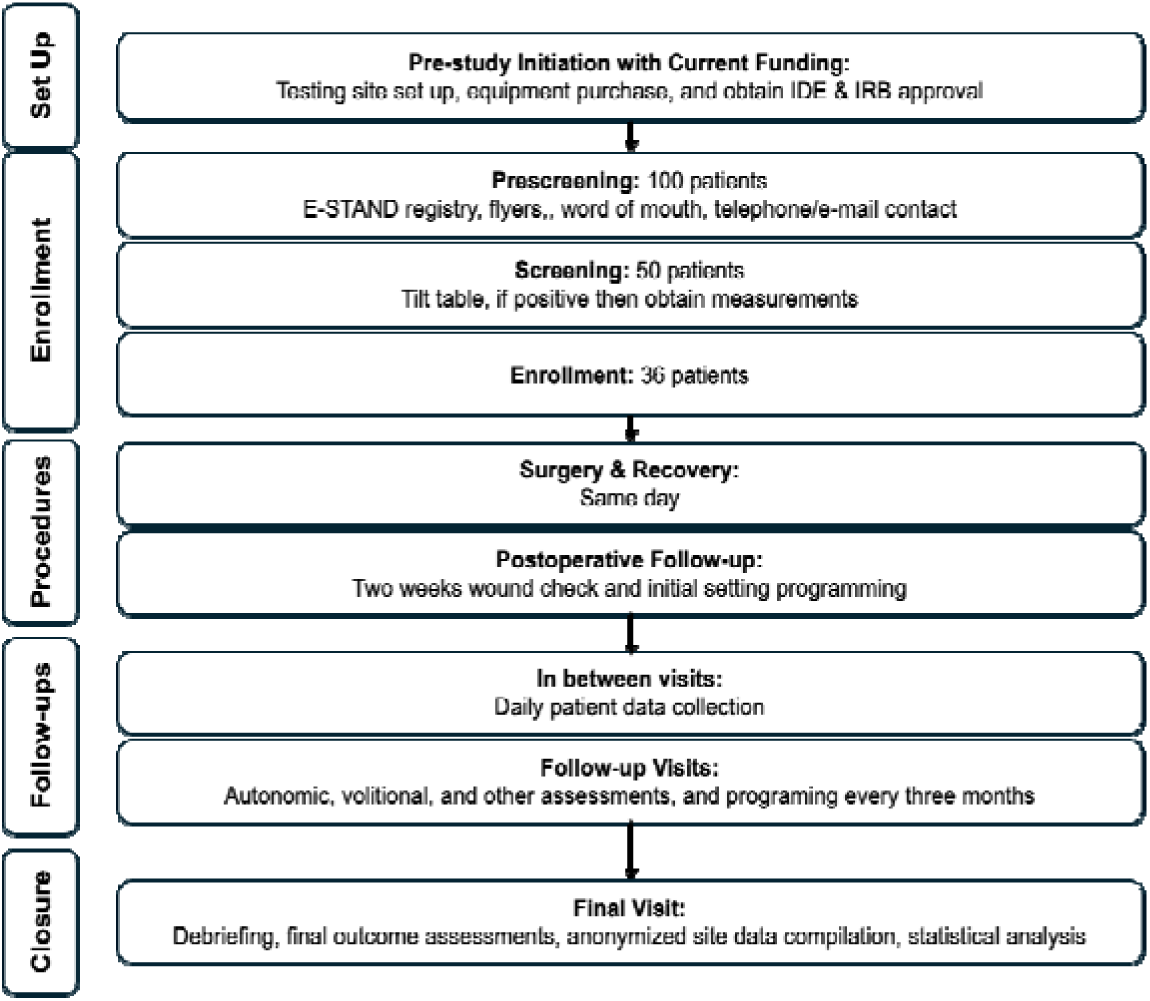
Overview of the CE-STAND trial.

After the first three participants are enrolled, three months of safety data will be acquired before enrolling the next participants. If there are no major safety issues, three additional participants will continue to be enrolled every three months (approximately one per month)), and the adverse events will be monitored throughout the entire follow-up period.

Therefore, our first milestone will occur at the six-month timepoint. We plan to have successfully enrolled three participants without significant safety events, with data from the first testing session. The team will review the data and adverse events (AEs) and determine whether any protocol modifications need to be made. We also plan to have the next three participants prepared for implantation, and an additional three scheduled for a consenting visit.

Our second milestone will occur at the end of the first year and annually thereafter. We will assess enrollment, review AEs, and evaluate study data.

Individuals with chronic SCI are highly motivated and very involved in their care. Retention strategies with include routine communication in which the study coordinator will contact the participants for monthly check-ins and to discuss any needs the participants may have. In addition, they will attend a testing session every 3 months. They will receive appointment reminders the week before their appointments and regular phone calls if their visit becomes delayed by more than one week.

### Study procedures

#### Screening Procedures

After consenting the participant, the study team member will schedule a separate appointment at the Cardiovascular and Rehabilitation laboratory to conduct the following:

⍰ Additional medical history will be reviewed as well as additional imaging if not previously reviewed (this may be reviewed prior to the screening visit)
⍰ Baseline medical examinations will be performed. Tilt table testing will be performed to confirm the presence of dysautonomia. Straps will be applied for safety and adjusted for comfort after screening for early signs of autonomic dysreflexia during preparation. The participant’s BP will be taken in supine position for 10 minutes and considered as the baseline. The tilt table will be moved initially to 30–45-degree and gradually increased to 60-70-degree tilt as tolerated, while having a BP measured continuously with a beat-to-beat non-invasive blood pressure monitor (NOVA, ADInstruments) and manual recordings as needed for accuracy. Participants will remain tilted for 10 min for continuous recordings and then placed in a supine position while BP is continuously measured for an additional 10 min.
⍰ A foot plate will not be used, as it could increase muscle activation and confound the assessment of orthostatic responses. Lower limb muscle activation will be recorded using non-invasive EMG and the modified Ashworth Scale to assess spasticity. If dysautonomia is not observed, then the participant will be deemed ineligible and no further testing will be performed.
⍰ If dysautonomia is observed, then further baseline testing will include tilt table testing with transcranial doppler (TCD) and concurrent N-Back cognitive exam.
⍰ Baseline modified brain motor control assessment (mBMCA), upper extremity testing, and modified functional reach test (mFRT) will also be performed. Additionally, baseline questionnaires will be administered.

#### Pre-Operative Visit

Study staff will verify inclusion/exclusion criteria and obtain baseline information according to the assessment schedule for registration purposes in the hospital clinical database. Further, participants will undergo:

⍰ Testing for cardiovascular autonomic function during the baseline will include: tilt table testing with BP and cardiac monitoring, TCD to assess cerebral blood flow, and N-Back cognitive testing, and microneurography to assess MSNA. To address limitations in isolating MSNA signals, we will record MSNA with brief bladder mechanical pressure^21^. Recordings will be conducted under controlled conditions using high-impedance, noise-shielded systems. They will be performed with stimulation off first to prevent contamination from the stimulator. Subsequently, we will measure the noise frequency and use filtering techniques to reduce the noise when the stimulators are on. Further, we will record MSNA from either the radial or peroneal nerves to increase the likelihood of obtaining a clear and stable signal.
⍰ Testing for volitional movement: This will include the mBMCA using non-invasive electromyogram (EMG). Further testing of upper extremities will be included with EMG, as well as the International Standards for Neurological Classification of SCI (ISNCSCI) documentation of upper extremity weakness, and the Graded Redefined Assessment of Strength, Sensibility, and Prehension (GRASSP) functional score.
⍰ Other testing: The mFRT will be performed to assess truncal stability.
⍰ Questionnaires will be completed for bowel and bladder function, sexual function, spasticity, pain, sleep, depression, and QoL.

Participants will be provided with a falls diary to maintain a log of fall events. Additional diaries will be provided to track participants’ BP, urinary and bowel function. Subsequently, preoperative and pre-anesthesia clinic assessments, along with pre-surgery education will be performed, and the surgery date will be scheduled.

#### Post-Operative Visit

A focused physical exam and wound inspection will be conducted two weeks post-operatively to ensure recovery from the procedure and assess for AEs such as infection. Study-specific AEs include hypotension, other hemodynamic instability, infection, bleeding, significant pain, or cerebrospinal fluid leak attributable to study participation. During the first 30 days, antiplatelet agents such as aspirin, or non-steroidal anti-inflammatory drugs (NSAIDs) such as ibuprofen may be withheld based on the clinical evaluation of each participant.

Initial stimulation settings will be programmed during the visit based on parameters shown to be effective in the E-STAND trial. These settings will be programmed individually for cardiovascular and volitional goals (eight and seven settings, respectively, totaling 15 settings, the maximum for this stimulator). For participants who demonstrate limited or no ability to use their hands, it may not be safe for them to adjust their settings independently. We will train the primary caregiver or personal support worker on device programming of the device for these participants. Subsequently, four follow-up visits will be scheduled.

#### In between visits

Participants will complete daily diaries that provide a binary comparison between the setting used that day and the previous day across multiple outcomes including BP, volitional movement, bowel and bladder function, as well as functional outcomes such as spasticity, posture, and transfers. A falls diary will be a participant-maintained log of fall events. Participants will be given setting assignments (one per day) at the previous visit, and these will be randomly assigned and participants will be blinded as to the reason for that particular setting choice.

Participants will have the ability to adjust stimulation parameters throughout the day according to their preferences.

They will be able to adjust the settings using a mobile app. Further, the Eterna stimulation system has a neuromodulation programming platform that enables investigators to monitor the device settings remotely.

Electronic questionnaires (e-questionnaires) will be sent to participants one week prior to their scheduled visit to complete at their own convenience before coming in. Participants will have the opportunity to complete the e-Questionnaires during visits if they couldn’t do so prior to the visit.

#### Follow Up Visits

The follow-up visits will take place every three months. During each visit the medical history and physical exam will be updated. Additionally, participants’ diaries and AEs will be reviewed. Participants will undergo the following testing procedures. The cardiovascular and volitional function will be assessed twice, once with stimulation on and another with stimulation off:

⍰ Testing for cardiovascular autonomic function will include: tilt table testing with BP monitoring, cardiac monitoring, TCD to assess cerebral blood flow, N-Back for cognitive testing, and MSNA.
⍰ Testing for volitional movement will include: the mBMCA using non-invasive EMG. Further testing of upper extremities will be included.
⍰ Other testing: The mFRT will be completed. Questionnaires will be completed for bladder, bowel, and sexual function, as well as QoL. The questionnaires will include assessments of:

#### Bowel and Bladder Function

The neurogenic bowel dysfunction score will be used to measure changes in bowel function and incontinence ^22^. The neurogenic bladder symptom score (NBSS) ^23^, the incontinence - QoL questionnaire ^24^, the autonomic dysreflexia health related quality of life (AD-HR QoL)^25,26^ questionnaire related to autonomic dysreflexia (AD) symptoms from bladder function and daily life, Penn Spasm Frequency Scale ^27^ and the Qualiveen questionnaire ^28^ will be used to assess changes in bladder function and incontinence.

#### Sexual Function

To assess sexual function, different metrics will be administered in the study. The Orgasm Rating Scale (ORS) will be used for men and women. Further, men will receive the International Index on Erectile Function questionnaire (IIEF) ^29^. Women will receive the Female Sexual Distress Scale questionnaire ^30–32^ and the Female Sexual Function Index (FSFI) questionnaire ^32–35^.

#### QoL

The QoL will be measured by the World Health Organization (WHO) QoL-BREF QoL Assessment tool ^36^. It is a 26 item questionnaire derived from the WHO-QoL 100, and the QoL Basic Data Set, which includes a 3-question summary questionnaire from the International SCI Data Sets ^37^.

#### Other Questionnaires

Spasticity will be assessed by the modified Ashworth score^38^ In addition, the Epworth Sleepiness Scale ^39,40^ will be used to determine the interference of drowsiness from SCI associated sleep disordered breathing in day-to-day activities.

For pain, the International SCI Pain Questionnaire Basic Data Set will be used to record and track the general pain profiles of all participants during the study ^41,42^. The participant will be asked about the type, location, and intensity of any pain issues they are having.

The Patient Health Questionnaire Depression Scale and Hospital Anxiety and Depression (PHQ-9)^43^ Scale will be administered. Positive psychiatric screening will result in a referral to the appropriate services. Positive suicidal/ homicidal ideation or intention to harm self or others will result in a referral to the appropriate services and immediate prompting of the proper authorities.

At the end of the visit, the stimulator will be programmed with 15 settings for the patient to test daily at home. All settings will be tested for safety and tolerability in the clinic to ensure that they do not induce any adverse hemodynamic responses. Participants will be instructed on self-monitoring for symptoms of autonomic dysreflexia, advised to immediately discontinue stimulation, and to document the event in their BP diary.

Real-time monitoring of BP and heart rate (HR) will be performed using continuous BP monitor during tilt table testing, whenever the stimulator is “on”, and during stimulator programing to detect autonomic dysreflexia and to capture both sympathetic activation and inhibition responses.

### Interventions

The spinal cord stimulator system (Abbott Eterna system implantable pulse generator (IPG), charger, and TriCentrus lead) will be used for this study which is manufactured by Abbott Laboratories. The surgical implantation of the system will be conducted by a neurosurgeon. The surgical procedure is similar to that for patients with chronic pain, which is a routine neurosurgical operation. First, EMG needles will be placed after general anesthesia is administered. Using conventional sterile technique and under fluoroscopic guidance, paddle electrodes will be inserted at the T10-T12 area to cover the lumbar spinal cord segments, through a small laminectomy. The desired targets of the electrodes will be T9-T12 for the superior two electrodes and T12-L2 for the inferior two electrodes ^12,44,45^.

To verify the coverage and placement of the eSCS paddles with suprathreshold stimulation, intraoperative neuromonitoring with EMG will be performed. In addition, electrophysiology mapping will verify the proper placement of the stimulator as detected by evoked motor responses with suprathreshold stimulation of the lumbar and upper sacral nerve root.

Subsequently, a subcutaneous pocket will be created to place the IPG. The paddle electrode wire will be tunneled subcutaneously to the pocket and connected to the neurostimulator ^12,16,17^. Participants will recover in the post-anesthesia care unit.

There are no plans to remove the device unless it malfunctions or complications/medical issues occur, requiring surgical removal of the device. Further, participants may continue to use the device after the end of the study unless they opt for it to be removed.

### Outcomes

The safety of eSCS usage in chronic cervical individuals with SCI is our primary outcome. The primary outcome will identify the incidence of AEs and serious adverse events (SAEs). Safety concerns, including hypotension, other hemodynamic instability (such as orthostatic hypotension with autonomic dysreflexia, cardiac arrhythmias, and shock), infection, bleeding, significant pain, spasticity, or cerebrospinal fluid leak attributable to study participation will be assessed post-operatively and during the follow-up visits. Additionally, safety concerns related to stimulation effects are particularly relevant in this patient population. We hypothesize that eSCS will be safe in individuals with cervical SCI.

For the secondary outcome, we will assess the efficacy of eSCS on autonomic function in individuals with chronic cervical tetraplegia, including cardiovascular and cognitive effects. These will include changes in BP and heart rate (HR) during tilt table testing; cognitive function using N-back testing; cerebral blood flow using TCD; and MSNA during microneurography. We hypothesize that epidural stimulation will increase resting SBP and CBF, decrease HR, and improve both cognitive function and MSNA compared to the sham condition.

For the tertiary outcome, we will evaluate the efficacy of eSCS on other functional modalities in chronic cervical individuals with SCI. These will include: Volitional function on EMG using the mBMCA and the volitional response index (VRI); upper and lower extremities using ISNCSCI and hand function using GRASSP functional score; truncal stability using the mFRT. We will administer questionnaire-based information to collect data related to bowel, bladder, and sexual function, spasticity, pain, sleep, depression, and QoL. We hypothesize that epidural stimulation will increase VRI magnitude and improve subjective outcomes across these domains compared to baseline or sham conditions, while hand function and truncal stability are expected to remain unchanged.

Each hypothesis will be tested using a two-tailed comparison, with the null hypothesis stating no difference between treatment and sham means for each measure.

Furthermore, to identify optimal stimulation settings for improved autonomic and volitional functional outcomes, Bayesian process and probit modelling stimulator settings will be utilized.

## Methods: Assignment of Intervention

### Allocation

During each clinical visit, participants will be tested for their preferred setting. A study member will program the settings into the stimulator. A schedule of device settings will be administered according to the stimulator optimization setting algorithm. Administration of stimulator settings will be randomized.

Accordingly, participants will be provided with a sequence of 15 settings every three months to test daily at home. Settings will be divided into patterns that target the lower segments (for volitional control n= 7), and higher segments (for autonomic effects n= 8) settings. With approximately 20 settings tested during these clinical visits, we will obtain a robust sampling of the parameter space and gain high confidence in identifying the optimal settings for blood pressure (BP) regulation and volitional movement separately.

### Blinding

The participant and clinician will be blinded to the stimulation setting to mitigate bias. However, blinding participants to when the stimulators are “on“ vs “off” will not be complete. Due to the significance of previously reported effects, participants will likely be able to recognize the difference between sham and treatment periods. Nonetheless, participants will be blinded to the stimulator’s programmed settings and to the specific settings used during study procedures when the stimulators are on.

## Methods: Data Collection, Management, and Analysis

The settings for each study participant will vary according to the experimental protocol. The pulse generator can deliver pulses of stimulation with varying amplitude (0-25.5 mA), frequency (2-1200 Hz), and pulse width (20-1000 μs). The amplitude is determined by the participant to adjust stimulation for comfort with the position. Optimization will occur during clinic visits after implantation.

### Data Management

Prior to each visit, setting preferences will be analyzed, and a response surface model will be fitted to the resulting data to extrapolate to the next series of settings that could potentially better inform the model, either by refining the parameter space near already preferred areas or by investigating unexplored regions.

We will optimize the stimulation parameters for each participant mainly by utilizing their responses to determine which stimulation patterns they prefer (cardiovascular and volitional). In addition to this, we will utilize the 24-hour BP paradigm utilizing their “favorite” setting with concurrent diary to determine whether their preferences for cardiovascular settings correlate with their actual BP readings.

Furthermore, we will conduct additional in-clinic testing to determine whether participant-selected settings are safe, tolerable, and effective for both cardiovascular and volitional testing and align with our objective findings. During, the clinical visits, vital signs and participant-reported symptoms will be monitored continuously. We will generate a model of the relationship between BP and parameters, and volitional movement and parameters (two separate processes), using that model to select new settings for testing. This process is called Bayesian Optimization ^46^. Settings with the highest preference will be retested to assess reproducibility.

### Statistical Methods

Participants who complete at least 75% of the protocol (3 out of 4 follow-up visits) will be included in the analysis dataset. Baseline characteristics will be described, including demographics, medical history, and all baseline assessments for this single-arm study. Descriptive statistics will be reported as means ± standard deviations. Tests will be considered statistically significant if alpha is less than 0.05 for two-tailed tests. All assumptions underlying statistical tests will be evaluated before use of the test and corrected if necessary and possible.

The repeated measures analysis of variance (ANOVA) will be used to compare responses during non-stimulation and stimulation, where alpha is assumed to be 0.025 (two tailed) and power is set at 0.95. The correlation between repeated measures is assumed to be 0.5. The ANOVA residuals will be assessed for normality and the groups will be assessed for homoscedasticity. Additionally, we will examine residuals and variance homogeneity to ensure the appropriateness of parametric analyses and report any deviations. If there are significant violations of these assumptions, Friedman’s test will be used as an alternative.

Additional sub-Group analyses will be completed with covariates of sex and age for the primary and secondary outcomes. Lesion location will also be used in a sub-group analysis for primary and secondary outcomes, but it is anticipated that there may not be sufficient variation for this analysis to be completed. The data will be displayed as means and 95% confidence intervals between stimulation and non-stimulation for each participant and as a group over time.

Missing data will be analyzed to examine for randomness of omission. If the missing data is determined to be reasonably random, then predictive mean matching will be used for imputation. The distribution of the complete data set will be examined with and without the imputed data. Data from subjects with incomplete data from dropout will be included in the final analysis. Incomplete data will be pooled per session

## Methods: Monitoring

### Auditing & Data Monitoring

A monitor has been assigned through the UMN Clinical and Translational Science Institute (CTSI) who is qualified by training and experience. Routine monitoring will occur to verify accuracy regarding enrollment, data collection, and AE monitoring and will report to the principal investigator (PI) and the local Institutional Review Board. The monitor will assure that the investigators comply with the signed investigator agreement, the Clinical Investigation Plan/Protocol, investigational device exemption (IDE) regulations and any conditions of approval imposed by the Investigation site’s IRB (Institutional Review Board)/IEC (Independent Ethics Committee) or Food and Drug Administration (FDA). Routine monitoring will occur to:

- Verify that participant enrollment is being achieved
- Verify that the inclusion/exclusion criteria has been met at enrollment
- Verify that the correct version of the informed consent has been signed by the participant
- Review the medical records of all enrolled participants to ensure all AEs have been captured and properly reported
- Verify that the data and imaging are accurate, complete, and backed up by source documents
- Verify that all contracts, certifications, and medical licenses for each site are valid through the duration of the study

The PI will monitor data and interpret the respective imaging. The investigator-sponsor will oversee and process data in the following manner:

- Data Management

- Design and modify data dictionary (the information that is being collected)
- Quality Control
- Retrieval and Reports

- Assist in preparation of data quality reports
- Assist in preparation of progress reports
- Design and Analysis

- Consult with biostatistician in design and analysis of research questions as needed

### Harms

The primary risks of participating in this study are related to surgical implantation of the spinal cord stimulators and testing. Risks due to surgery are related to undergoing general anesthesia, surgery, and malfunction of the device itself. Participants will undergo anesthesia for implantation which harbors risk including heart arrhythmia, pneumonia as well as rare risks such as heart attack, stroke, and death. Surgical risks are well-known and documented in the literature. Additional surgery may be required if the device malfunctions, an infection develops, or cerebrospinal fluid leak results.

In our existing study of eSCS after motor-complete SCI, we have not observed infections associated with the intervention or any serious adverse outcome. Hardware stimulator failure has been observed twice due to the significantly higher requirements for stimulation for SCI, but no adverse outcome has occurred. Typically, participants in the study have had the same or reduced rates of infection, falls, and injury associated with SCI.

## Ethics and Dissemination

### Research Ethics Approval

FDA approval of the study protocol was obtained using an IDE for the Abbott Eterna system IPG and TriCentrus lead. Subsequently, the study was approved by the UMN IRB. It is registered on ClinicalTrials.gov (NCT06410001).

### Protocol Amendments

Site-specific protocol amendments are available upon request from the corresponding author.

### Consent

When the PI or PI-delegated study personnel identify a potentially eligible candidate, the delegated study personnel will schedule an appointment to obtain consent. The consent process will take place electronically, utilizing the REDCap e-consent platform. Participants will be provided with secure access to the e-consent form before their appointment. This process will ensure compliance with institutional and regulatory standards for electronic informed consent.

The REDCap e-consent will take place at the study site with both the PI-delegated study personnel and the participant present, or remotely when the participant is at home or another convenient and private location. When done remotely, the participant will discuss the e-consent document with a person authorized to obtain study consent; this discussion often occurs via phone or video conference. For participants with diminished or limited capacity, the IRB may require a video connection during the consent conversation.

### Confidentiality

All records related to a participant’s research will be stored in locked filing cabinets with access restricted to study personnel or on password-protected computers in compliance with HIPAA standards. The participant’s identity on these records will be indicated by a study number rather than by name, and the information linking these study identification numbers with the participant’s identity will be kept in a separate file on Box. The participant will not be identified by name in any publication of the research results. Participant data will be de-identified and linked to unique study identifiers. Data quality and integrity will be maintained through regular monitoring and inventory checks. For long-term preservation and sharing study data, protocol, and metadata will be deposited in DRUM.

### Access to Data

The collection, transfer, and storage of data will be conducted in compliance with the HIPAA Security Rule and structured to minimize the risk of protected health information (PHI) disclosure. All recorded data will be entered into a password-protected database. All data entry forms will be accessed and completed electronically through a password-protected log-in.

All user interactions with the web-based system, from transmitting access passwords to entering sensitive participant data, are done via 128-bit encryption using the secure HyperText Transfer Protocol Secure (HTTPS) protocol. Firewalls ensure that only the minimum traffic required for normal operations is allowed to traverse the network of web and database servers. The database production servers will be housed in secure institutional data center facilities and will include failover protection designed to minimize the potential for server downtime. Only the minimal required personnel are allowed direct access to production facilities. The study data will remain secure and be maximally protected.

Physical copies of participant research records will be secured by study personnel after review or use in locked file cabinets. Inventory of paper files by study personnel will occur monthly, and a checklist will be maintained to keep track of potential missing documents. Contact information for the study, including the name of the study, name of the PI, address of the study office, and phone number of the study will be added to study documents and study document containers in the unlikely event that study documents are misplaced.

Participants will be voluntarily sending us their medical records after which we will create an EPIC record for them in our system. Participants will have signed the study HIPAA authorization to allow researchers access to medical records. Participants will be given a unique identifier when they are determined to be eligible for preliminary screening.

Only the study investigators and study team member will have access to these documents. A copy of the participants’ signed consent form will be uploaded onto their EPIC file so their care providers are aware that they are part of an interventional trial that has been deemed to have a more than significant risk to participants. The PI will ensure the protection of participants’ privacy and will evaluate privacy protection at least once per year. The informed consent process and all study-related visits will take place in private rooms located in the Clinical Research Unit.

### Ancillary and Post-trial Care

The data collected from this clinical trial will be stored in Box and RedCap. Box Secure Storage is a secure environment delivered by the Center of Excellence for HIPAA Data. It is intended for storing, sharing, and accessing sensitive and highly restricted files. Box Secure Storage includes encryption, activity logging, Duo Two-Factor Authentication, and access controls such as view-only access. It is compliant with HIPAA and security standards for private, highly restricted data. A long-term data sharing and preservation plan will be used to store and make publicly accessible the de-identified data, including experimental results and protocols, beyond the life of the project.

The data will be deposited into the Data Repository for the University of Minnesota (DRUM). This University Libraries-hosted institutional data repository is an open-access platform for dissemination and archiving of university research data. Data files in DRUM are written to an Isilon storage system with two copies, one local to each of the two geographically separated UMN Data Centers. The local Isilon cluster stores the data in such a way that the data can survive the loss of any two disks or any one node of the cluster. Within two hours of the initial write, data replication to the second Isilon cluster commences.

The second cluster employs the same protections as the local cluster, and both verify with a checksum procedure that data has not altered on write. In addition, DRUM provides long-term preservation of digital data files for at least 10 years using services such as migration (limited format types), secure backup, bit-level checksums, and maintains a persistent Digital Object Identifier (DOI) for datasets, facilitating data citations. In accordance with DRUM policies, the (de-identified, if applicable) data will be accompanied by the appropriate documentation, metadata, and code to facilitate reuse and provide the potential for interoperability with similar datasets.

In the event of a potential breach of data confidentiality, the incident will be reported to the on-site IRB office within 5 business days of discovering the event. The study team will follow recommendations by the institutional IRB and privacy officer. Study personnel on site will make a detailed report and thoroughly investigate the possibility of file misplacement. Contact information will be maintained (in a secure, limited-access research drive) for all study participants to prepare for post-event communication and coordination in the event of a data breach for the duration of record retention.

### Dissemination Policy

This publication was prepared using the Standard Protocol Items: Recommendations for Interventional Trials (SPIRIT) ^47^ statement. The SPIRIT checklist can be found in Supplemental Additional File 1. We will perform an interim analysis to provide interim findings, and these will inform further research and address any issues in the study. These reports will be made available through peer-reviewed journals, conference presentations, and clinical network reporting.

### Patient and Public Involvement

The initial startup of this project was partially funded by the Minnesota Office of Higher Education (MN OHE) and included members of the community with lived experience. In addition, annual and quarterly reports are provided to update the MN OHE and Spinal Cord Society (SCS) Foundations’ members on the status of the study. Once the study is launched, patients and the broader SCI community will be involved in recruitment efforts and in the dissemination of study results.

## Supporting information

https://www.spirit-statement.org/wp-content/uploads/2013/01/SPIRIT-Checklist-download-8Jan13.pdf

## Data Availability

All data produced in the present study are available upon reasonable request to the authors

## Authors’ Contributions

Ann Parr / AP is responsible for the overall content as guarantor.

NM: Contributed to conceptualization of the study protocol drafting, and refinement.

MK: Contributed to study methodology, specifically for autonomic testing.

WL: Provided expertise and assisted in drafting and revising the manuscript.

DB: Assisted with study procedures and reviewed the manuscript for accuracy and clarity.

## Appendices

SPIRIT Checklist (supplemental additional file 1).

